# Epstein Barr viral shedding dynamics in saliva during acute SARS-CoV-2 infection

**DOI:** 10.64898/2025.12.31.25343091

**Authors:** Miguel A. Garcia-Knight, Jessica Y. Chen, Amethyst Zhang, Michel Tassetto, Scott Lu, Isidro X Perez-Añorve, Sarah A. Goldberg, Khamal Anglin, Badri Viswanathan, Steven G. Deeks, Timothy Henrich, Jeffrey N. Martin, Michael J. Peluso, Raul Andino, J. Daniel Kelly

**Affiliations:** Departamento de Inmunología, Instituto de Investigaciones Biomédicas, Universidad Nacional Autónoma de México, Mexico City, Mexico; Wellcome Sanger Institute, Hinxton, United Kingdom; Department of Immunology and Microbiology, University of California San Francisco, San Francisco, USA; Department of Epidemiology and Biostatistics, University of California, San Francisco, San Francisco, California, USA; Departamento de Ciencias Naturales, Unidad Cuajimalpa, Universidad Autonoma Metropolitana, 05348 Mexico City, Mexico; Division of HIV, Infectious Diseases and Global Medicine, Zuckerberg San Francisco General Hospital, San Francisco, California, USA; Division of Experimental Medicine, Department of Medicine, University of California San Francisco, San Francisco, CA 94143, USA; San Francisco VA Medical Center, Department of Medicine, University of California, San Francisco, San Francisco, California, USA; Institute for Global Health Sciences, University of California, San Francisco, San Francisco, California, USA; F.I. Proctor Foundation, University of California, San Francisco, San Francisco, California, USA

**Keywords:** SARS-CoV-2, Epstein-Barr virus, viral shedding, co-infection

## Abstract

**Background:** Epstein–Barr virus (EBV) establishes latency in most adults and can be reactivated under conditions of co-infection and immune dysregulation. COVID-19 has been associated with EBV reactivation, primarily in hospitalized cohorts, but EBV shedding in the oral cavity and the extent to which these dynamics trigger a systemic anti-EBV antibody response during acute COVID-19 remain poorly understood.

**Methods:** We conducted a nested cohort study of 69 community-based participants including 56 SARS-CoV-2–positive individuals and 13 uninfected household contacts, enrolled as part of a prospective cohort within 5 days of COVID-19 symptom onset. Participants self-collected saliva and nasal samples daily through day 14, then every 3 days until day 21, and once more at day 28. Blood samples were collected on day 28. SARS-CoV-2 RNA in the saliva and nares and EBV DNA in the saliva were quantified by RT-qPCR and qPCR, respectively. EBV antibody responses against viral capsid antigen IgM and IgG, early (D) antigen IgG, and nuclear antigen IgG were quantified in serum by ELISA.

**Results:** SARS-CoV-2–positive participants tended to be more likely to experience consecutive days of EBV shedding in saliva (*p* = 0.10) and contributed a higher mean number of EBV DNA–positive specimens compared with uninfected controls (5 vs. 1 per participant in EBV+ participants; *p* = 0.018). Among SARS-CoV-2–positive participants with EBV reactivation, a positive correlation was observed between maximum EBV DNA in saliva and SARS-CoV-2 RNA levels in saliva (*r* = 0.49, *p* = 0.047) but not with SARS-CoV-2 RNA levels in nasal samples (*r* = -0.12, *p* = 0.67). EBV reactivation in saliva was not associated with induction of serological responses to systemic EBV reactivation.

**Conclusions:** This cohort study provides evidence that SARS-CoV-2 infection is associated with prolonged salivary EBV shedding, that concurrent high levels of EBV and SARS-CoV-2 in saliva are correlated, but these salivary EBV shedding dynamics do not drive systemic EBV antibody responses. Further research with larger cohorts and mechanistic assays would be helpful to elucidate the biological pathway triggering systemic EBV antibody responses.

## INTRODUCTION

Epstein–Barr virus (EBV) is a ubiquitous human gammaherpesvirus, with approximately 90% of adults worldwide exhibiting serological evidence of prior infection(1). Following primary infection, EBV establishes lifelong latency, primarily within B lymphocytes, but retains the capacity for periodic reactivation (1). This process can be triggered by immune dysregulation, inflammation, or co-infection, leading to lytic replication and viral shedding that can be detected in blood or saliva. In the latter, reactivation of EBV is thought to initiate in B cells and to be amplified in epithelial cells of the oral cavity, including the nasopharynx and tonsils in a process that is highly dynamic and variable over time within individuals(2). Although EBV reactivation is often clinically silent, it has been implicated in a wide spectrum of pathological conditions, particularly in settings of impaired immune surveillance(3). The mechanisms by which EBV lytic activity is influenced by local mucosal factors and how this in turn impacts reactivation at the systemic level and influences pathology, remains poorly defined.

Coronavirus disease 2019 (COVID-19) is caused by severe acute respiratory syndrome coronavirus 2 (SARS-CoV-2), a respiratory pathogen with broad tissue tropism that, amongst many cell types, infects mucosal and salivary epithelial cells in the oral cavity. Infection is characterized by profound inflammatory responses, transient lymphopenia, and functional impairment of T cell–mediated immunity (4). These immune perturbations have been proposed as potential drivers of herpesvirus reactivation. Indeed, multiple studies have reported evidence of EBV reactivation in patients with COVID-19, particularly among hospitalized and critically ill cohorts, where EBV DNA has frequently been detected in plasma or whole blood. In such settings, EBV reactivation has been associated with increased disease severity, prolonged hospitalization, and higher mortality (5–7).

Despite these observations, important gaps remain in our understanding of EBV reactivation during SARS-CoV-2 infection. Most prior studies have focused on severe or hospitalized cases, limiting generalizability to community-managed infections(5–7). Additionally, EBV reactivation has been largely assessed using blood-based markers, which may not accurately reflect localized viral dynamics within mucosal compartments. Moreover, few studies have incorporated robust longitudinal samplings to examine within-host viral shedding patterns, and it remains unclear whether local EBV reactivation in saliva during acute COVID-19 is accompanied by systemic serological evidence of EBV reactivation(8, 9).

To address these gaps, we conducted a household-ascertained prospective cohort study of SARS-CoV-2–infected individuals and uninfected household controls. We quantified EBV DNA in saliva alongside SARS-CoV-2 RNA in both saliva and nasal specimens over the course of acute infection. Additionally, we evaluated antibody responses at convalescence targeting EBV antigens in serum of systemic reactivation. By focusing on the oral cavity, a site of EBV and SARS-CoV-2 co-infection, this work provides new insight into localized viral interactions during acute SARS-CoV-2 infection in a non-hospitalized population.

## METHODS

### Ethics

The study was reviewed by the University of California, San Francisco (UCSF) Institutional Review Board and given a designation of public health surveillance according to federal regulations as summarized in 45 Code of Federal Regulations 46.102(d)(1)(2). This activity was reviewed by Centers for Disease Control and Prevention (CDC) and was conducted consistent with applicable federal law and CDC policy.^§^ Written informed consent was obtained from all participants (IRB#20-30388).

### Participants

This was a nested cohort study within an observational, prospective, non-hospitalized case-ascertained household cohort from the San Francisco Bay Area designed to study the natural history of SARS-CoV-2 infection. Enrollment occurred from September 2020 until August 2023. Individuals from testing sites affiliated with the University of California, San Francsico, and San Mateo County Health Department with lab-confirmed SARS-CoV-2 (index cases) were enrolled within 5 days of symptom onset, and their household members were screened and recruited. Participants self-collected anterior nasal swabs and saliva daily for 14 days post-symptom onset (PSO), then every 3 days through day 21 PSO, and at day 28 PSO. Blood samples were collected weekly through day 28. During these weekly visits, staff also administered questionnaires. Additional details on the methods of the parent study can be found elsewhere (10, 11). This analysis was restricted to a subset of individuals of all ages who tested positive for SARS-CoV-2 and household contacts who remained SARS-CoV-2 negative for the follow-up period and for whom saliva and blood samples for EBV DNA and anti-EBV antibody testing were available.

### Questionnaire-based measurements

Questionnaires were used to collect data on the following covariates: age, sex, race/ethnicity, body-mass index, medical conditions, COVID-19 vaccination status (verified by vaccination card), and receipt of SARS-CoV-2 antiviral therapy.

### SARS-CoV-2 RNA quantification

SARS-CoV-2 RNA in the saliva and nares were quantified by RT-qPCR and targeted nucleocapsid (N) and envelope (E) genes on a CFX Connect Real-Time PCR detection system (Biorad). Details on this laboratory assay can be found elsewhere (11). Prior work showed a correlation between N and E genes, so this study reports SARS-CoV-2 RNA quantification targeting the N gene for simplicity.

### EBV DNA quantification

EBV DNA was extracted from 200uL of saliva mixed 1:1 with viral transport media composed of Hank’s Balanced Salt Solution (HBSS) with calcium and magnesium, 2% foetal bovine serum, 100 ug/mL gentamicin and 0.5ug /mL amphotericin B (Bio-world). The MagMAX Viral/Pathogen II Nucleic Acid Isolation Kit (Thermo Scientific) was used according to the manufacturer’s instructions, and the samples were processed on the Kingfisher (Thermo Scientific) automatic extraction instrument. The Epstein-Barr Virus (EBV) Real Time PCR Assay primer/probe mix (Virusys Corporation. Taneytown, MD, USA) was used according to the manufacturer’s instructions and run on the CFX Connect Real-Time PCR detection system (Biorad). The assay uses a primer and probe (FAM/BHQ-label) mix specific for BALF5 and an EBV standard with a linear range 1x10^8^ to 1x10^2^. To account for the presence of cellular EBV DNA in the saliva in the general population, a sample was considered EBV DNA positive if it was found to contain 500 EBV target copies/reaction or more (2, 12).

### EBV serology

Blood serum collected on day 28 of the study was used to test a panel of EBV antibody responses (ARUP Laboratories). The panel generated quantitative measures (units per mL) of viral capsid antigen (VCA) IgM and IgG, early antigen (EA)-D IgG, and nuclear antigen (EBNA) IgG. Results were considered positive in this analysis if units (U) per mL were higher than the indeterminate range of the assay (VCA IgM ≥44 U/mL; VCA IgG ≥22 U/mL; EA-D ≥11 U/mL; EBNA IgG ≥22 U/mL). The VCA IgG, EBNA IgG, and EA-D IgG assays had upper limits of quantitation (>750 U/mL, >600 U/mL, and >150 U/mL, respectively).

### Statistical analysis

Baseline characteristics were described for SARS-CoV-2-infected and -uninfected participants. A graphical depiction of SARS-CoV-2 RNA levels (log10 copies/ mL) in the saliva and nares as well as EBV DNA levels (log10 copies/ mL) in the saliva was longitudinally generated over the 28-day study period for each SARS-CoV-2-infected and -uninfected participant. EBV DNA positive results were defined as having at least one measurement above the cutoff of 500 copies/reaction. EBV-positive individuals were further defined as having consecutive positive results if more than one measurement over consecutive days was above the cutoff for EBV positivity. The frequency of individuals with any EBV positive saliva sample and the frequency of individuals with saliva samples positive for EBV over consecutive days was compared between SARS-CoV-2 infection groups using a Fischer’s Exact test. Continuous data was compared between groups using a Wilcoxon ranked sum tests. To account for zero-skewed data, comparisons were carried out on the total dataset and on non-zero values and both p values shown. Among the SARS-CoV-2 infected participants, maximum EBV DNA levels were correlated with maximum SARS-CoV-2 levels using Pearson correlation statistics. Statistical tests estimating a p-value of 0.05 or less were considered to be significant. Analyses were performed using R version 4.5.1 (R Project for Statistical Computing, Vienna, Austria).

## RESULTS

### Study participants

A total of 69 participants were included, 56 SARS-CoV-2–positive and 13 SARS-CoV-2–uninfected controls. Baseline demographics were largely comparable, though a higher proportion of SARS-CoV-2 uninfected participants were <30 years of age and a higher proportion of SARS-CoV-2 infected participants had received booster and bivalent booster vaccinations. A similar number of longitudinal specimens were analyzed in both groups. Amongst SARS-CoV-2 infected participants, 7/56 (13%) received Paxlovid and 32/56 (58%) were infected with pre-Delta or Delta variants, and the remainder with distinct lineages of the Omicron variant.

**Table 1:** Participant demographics, vaccination status, and EBV PCR results.

| Parameter | SARS-CoV-2 Uninfected<br>N = 13<sup>1</sup> | SARS-CoV-2 Infected<br>N = 56<sup>1</sup> |
| --- | --- | --- |
| Age (years), median (IQR) | 29.0 (19.0-43.0) | 41.0 (33.0-56.5) |
| Age (years), n (%) |  |  |
| <18 | 3 (23%) | 2 (3.6%) |
| 18-29 | 4 (31%) | 7 (13%) |
| 30-39 | 2 (15%) | 13 (23%) |
| 40-49 | 2 (15%) | 16 (29%) |
| 50-59 | 1 (7.7%) | 4 (7.1%) |
| >60 | 1 (7.7%) | 14 (25%) |
| Sex, n (%) |  |  |
| Female | 4 (31%) | 25 (45%) |
| Male | 9 (69%) | 31 (55%) |
| **BMI, median (IQR)** | 21.1 (18.5-25.1) | 25.8 (23.2-29.5)* |
| **Vaccination status, n (%)** |  |  |
| Unvaccinated | 3 (23%) | 20 (36%) |
| One dose | 3 (23%) | 1 (1.8%) |
| Primary series | 6 (46%) | 15 (27%) |
| Boosted | 1 (7.7%) | 11 (20%) |
| Bivalent booster | 0 (0%) | 9 (16%) |
| **Specimens analyzed/participant, median (IQR)** | 13.0 (13.0-14.0) | 14.0 (13.0-15.0) |
| **Received Paxlovid, n (%)** | NA | 7 (13%) |
| **SARS-CoV-2 variant** |  |  |
| PreDelta | NA | 21 (38%) |
| Delta | NA | 11 (20%) |
| BA.1 | NA | 4 (7.1%) |
| BA.2 | NA | 2 (3.6%) |
| BA.5 | NA | 10 (18%) |
| XBB.1.5 | NA | 8 (14%) |

\*Data from one individual not collected

### EBV shedding over consecutive days in saliva during acute SARS-CoV-2 infection

We evaluated EBV shedding dynamics in saliva over 28 days in SARS-CoV-2 infected and uninfected individuals, respectively (Fig. 1A&B). In the same individuals, we also evaluated SARS-CoV-2 shedding in nasal and saliva specimens. We found no difference in the proportion of SARS-CoV-2-infected and uninfected individuals with any detectable EBV DNA over the study period (4/13, 30.8% vs 17/56, 30.4%). However, the proportion of individuals with consecutive days of EBV shedding tended to be higher in SARS-CoV-2 infected individuals compared to controls (0/13, 0%, vs 13/56, 23.2%, P=0.10); although the p-value was not statistically significant, only SARS-CoV-2 infected individuals were observed to exhibit EBV shedding over consecutive days. Notably, longitudinal shedding patterns of EBV and SARS-CoV-2 were mirrored, particularly in saliva, in individuals 4527 and 4789 (Fig. 1B), with the latter having received Paxlovid treatment and clearly experiencing SARS-CoV-2 viral rebound(13). Together, these data suggest that whilst EBV was sporadically detectable in uninfected individuals, sustained EBV shedding in saliva over consecutive days may be more common in the context of SARS-CoV-2 infection.

**Figure 1.**
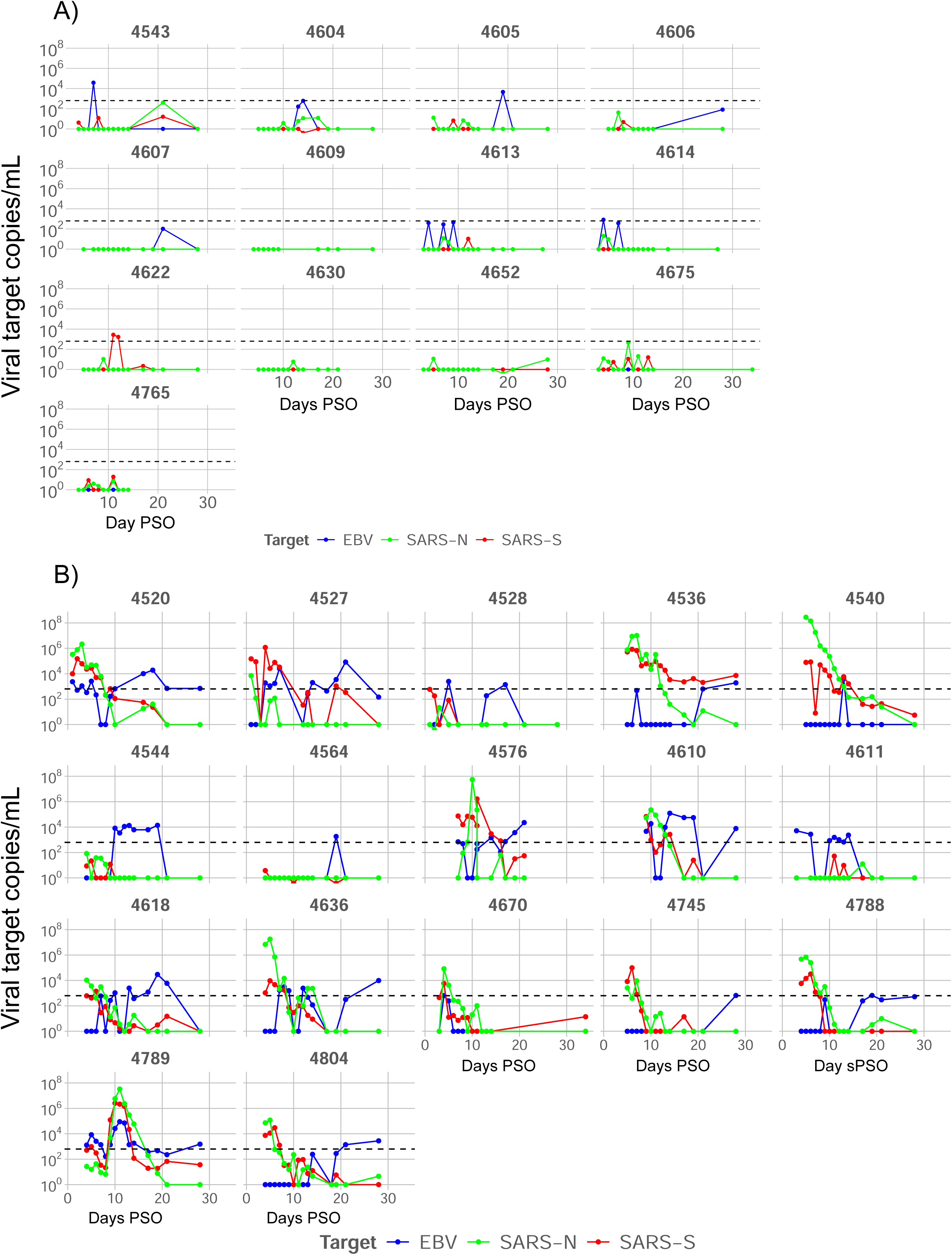
Temporal SARS-CoV-2 and EBV shedding dynamics in SARS-CoV-2 infected and uninfected individuals. A) Individual viral shedding dynamics over 28 days from enrollment in all SARS-CoV-2 uninfected individuals included in the study; B) Individual viral shedding dynamics over 28 days in SARS-CoV-2 infected individuals from the day of symptom onset. Only participants with detectable EBV above a threshold of 500 viral DNA copies/mL are shown. Colored lines represent SARS-CoV-2 RNA copies/mL in saliva and nasal swabs and EBV DNA copies/mL in saliva, as indicated. PSO, post symptom onset.

### Prolonged EBV shedding in SARS- CoV-2 infection

To further assess differences in EBV reactivation between groups, the number of EBV-positive samples per participant and the proportion of EBV-positive specimens per participant were compared. Individuals with SARS-CoV-2 infection had a significantly greater number (Fig. 2A, P=0.018) and proportion (Fig. 2B, P=0.03) of specimens that were EBV positive during the follow-up period compared to SARS-CoV-2-uninfected controls. In contrast, no differences were observed between the maximum EBV viral load between groups (Fig. 2C). This finding further supports the observation that EBV reactivation was more prolonged during SARS-CoV-2 infection, though this is not associated with maximum EBV viral loads in saliva.

**Figure 2.**
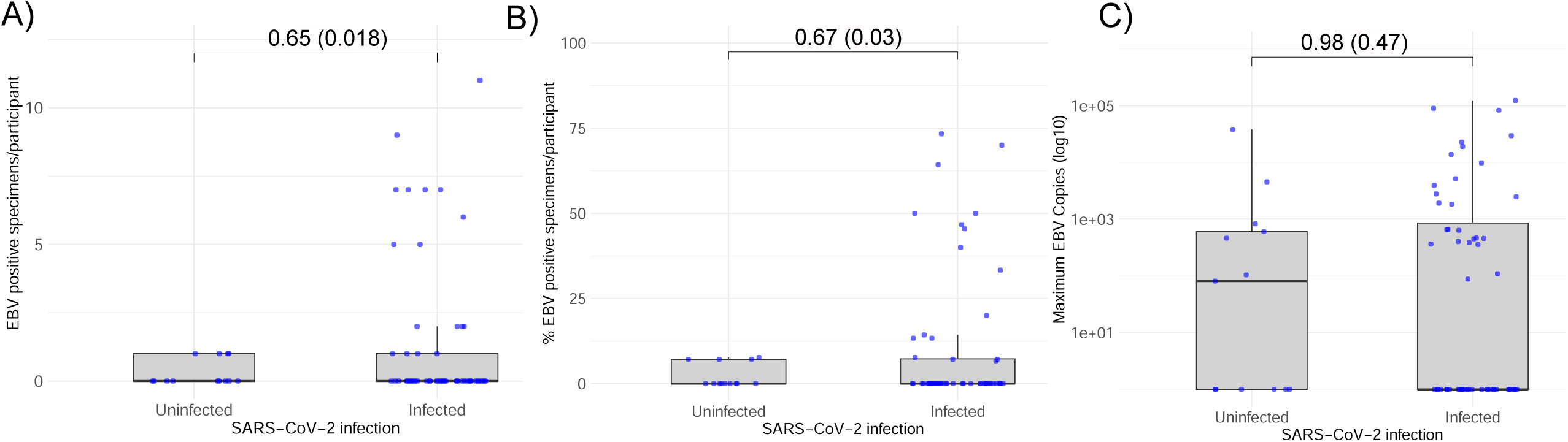
Comparison of EBV shedding in SARS- CoV-2 infected and uninfected individuals. A) A box plot comparing the number of EBV-positive saliva samples in SARS-CoV-2 infected and uninfected individuals. (B) A box plot comparing maximum EBV DNA copies in longitudinal saliva samples from SARS- CoV-2 infected and uninfected individuals. To account for zero-skewed data, Wilcoxon ranked sum tests were carried out on the total dataset and on non-zero values. Adjusted P values for non-zero values are shown in parenthesis.

### Maximum EBV viral load correlated with maximum SARS-CoV-2

As SARS-CoV-2 infection was associated with EBV shedding in saliva, we assessed, amongst SARS-CoV-2 infected individuals, whether there was a quantitative relationship between the viral loads of both infections. Maximum EBV DNA copies in saliva correlated positively with maximum SARS-CoV-2 RNA copies in saliva (r=0.49, P=0.047; Fig 3A) but not with those in nasal specimens measured over the study period (r=-0.12, P=0.67; Fig 3B).

**Figure 3.**
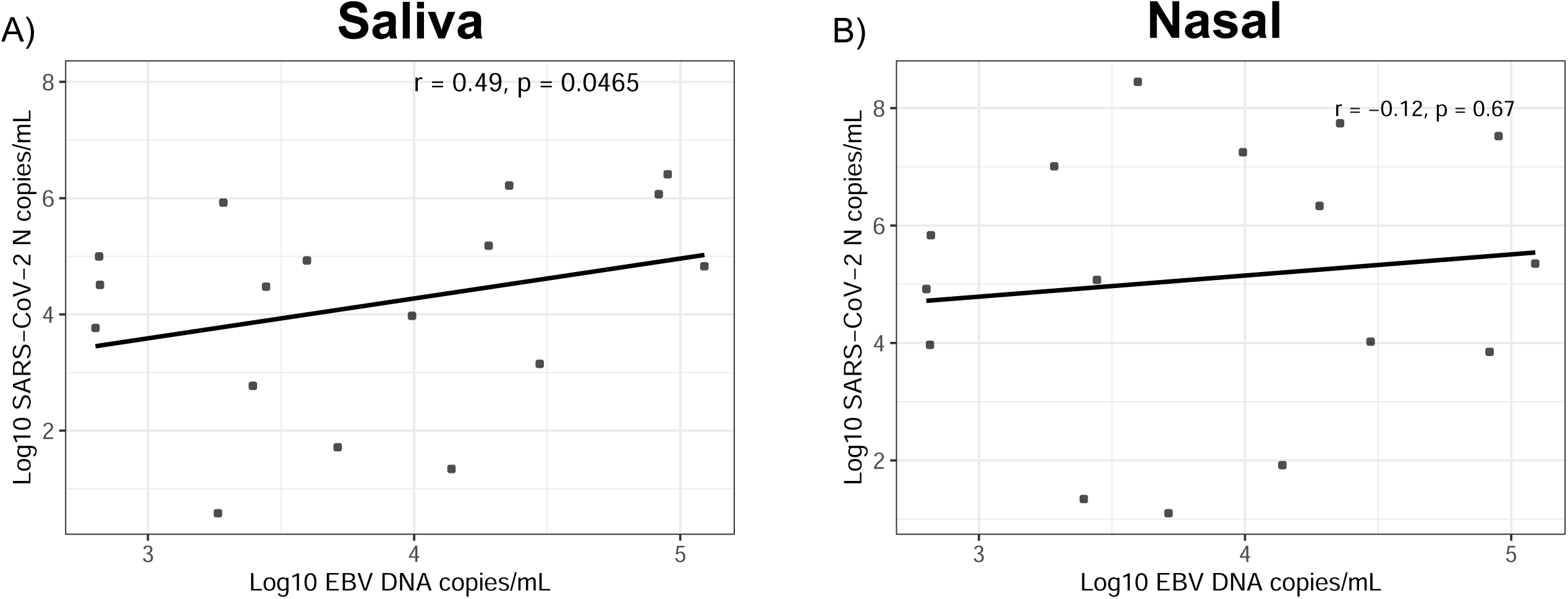
Maximum SARS-CoV-2 and EBV viral copies in SARS-CoV-2 infected individuals with EBV reactivation. A) Correlation between the maximum copies of SARS-CoV-2 nucleocapsid (N) RNA in saliva (log10 copies/mL) and the maximum copies of EBV DNA in saliva (log10 copies/mL) over the study period. B) Correlation between the maximum copies of SARS-CoV-2 nucleocapsid (N) RNA in nasal specimens (log10 copies/mL) and the maximum copies of EBV DNA in saliva (log10 copies/mL) over the study period. The analysis was restricted to SARS-CoV-2 infected participants with detectable EBV DNAemia in saliva above a threshold of 500 viral DNA copies/mL. Pearson correlation coefficient and P value are shown.

### EBV DNA shedding in saliva did not correlate with systemic EBV antibody responses at day 28 following SARS-CoV-2 infection

We next assessed whether EBV shedding in saliva over consecutive days observed following SARS-CoV-2 infection was in turn associated with a serological response against systemic EBV reactivation. For this, we focused on proposed surrogates for systemic EBV reactivation, including viral capsid antigen (VCA) IgM and early antigen diffuse (EA-D) IgG(8). We also measured EBV nuclear antigen [EBNA] IgG and VCA IgG, responses that are sustained throughout chronic infection, with the titres of the former having been shown to increase during lytic reactivation(14). Antibody responses to these antigens were measured in plasma in 45/56 (80.4%) of SARS-CoV-2 infected individuals with available serum specimens 28 days after SARS-CoV-2 infection. We observed significantly increased levels of EA-D IgG in SARS-CoV-2 infected participants without EBV reactivation in saliva compared to those with reactivation (Figure 4A, P=0.021). No significant differences between these groups were observed with regards to VCA IgM, EBNA IgG or VCA IgG (Figure 4B-D), and no association was found between detection of EBV DNA in saliva over consecutive days following SARS-CoV-2 infection and serological markers of EBV reactivation (Figure 4A-D, red dots).

**Figure 4.**
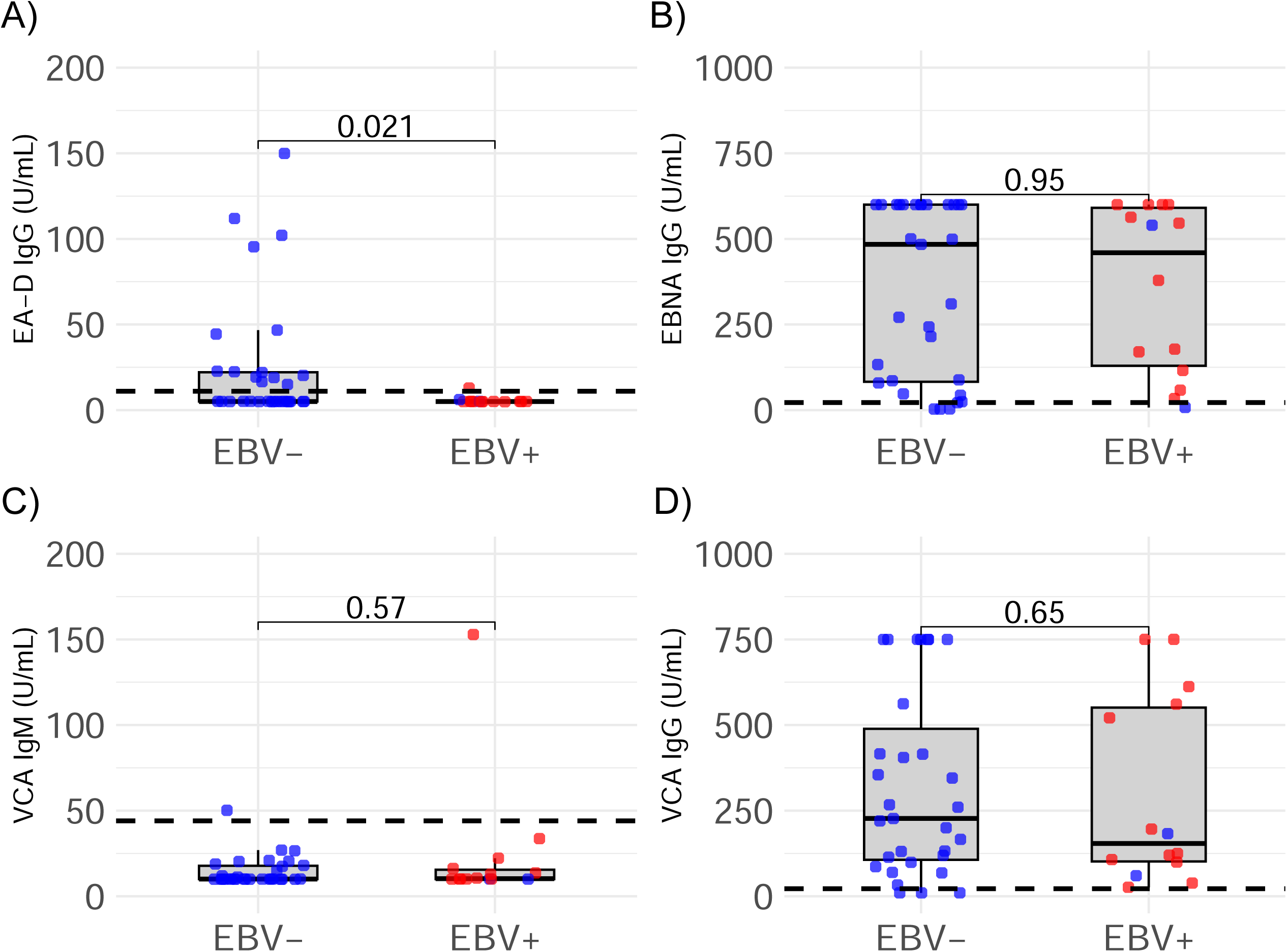
Comparison of EBV serological response amongst SARS-CoV-2 infected participants with and without EBV reactivation in saliva. Antibody titers of early antigen diffuse (EA-D) IgG (A), nuclear antigen (EBNA) IgG (B), viral capsid antigen (VCA) IgM (C) and VCA IgG (D). Antibody titers were compared between EBV shedding groups. Assay cut offs are indicated by a dashed line. Participants that had EBV positive saliva specimens on two or more consecutive days are shown in red and those with no EBV positive saliva samples on consecutive days are shown in blue. Wilcoxon ranked sum tests were carried out to assess differences between groups and P values are shown.

## DISCUSSION

In this study of a household-ascertained prospective cohort, we observed sustained EBV DNA shedding over consecutive days in close to a quarter of SARS-CoV-2-infected individuals, a feature absent from uninfected controls. Furthermore, among individuals with detectable EBV reactivation, we observed a positive correlation between maximum EBV DNA levels and maximum SARS-CoV-2 RNA levels in saliva, but not in nasal specimens. By contrast, EBV salivary shedding was not associated with serological markers of systemic EBV reactivation measured during convalescence. Together, these findings suggest that SARS-CoV-2 infection promotes localized EBV reactivation within the oral cavity, while such mucosal viral activity may remain largely uncoupled from the processes that trigger a systemic response to EBV reactivation.

Previous studies examining EBV reactivation in the context of COVID-19 have primarily focused on hospitalized or critically ill patients and have relied on detection of EBV DNA in blood. In these settings, EBV reactivation has been reported in up to 80% of patients and has been linked to adverse clinical outcomes, including prolonged intensive care unit stays and increased mortality (5). Our findings extend these observations by demonstrating that EBV reactivation can also occur in a sustained manner in saliva in non-hospitalized SARS-CoV-2 infections. However, a longitudinal study design with dense within-host sampling of saliva was critical for identifying these changes, as we found that the proportion of individuals with any EBV positive saliva specimens over the study period was not modulated by SARS-CoV-2 infection. Accordingly, a cross-sectional study of acute COVID-19 cases observed no differences in EBV shedding in saliva between cases and controls(15).

The observed correlation between EBV and SARS-CoV-2 viral loads within saliva points to a potential role for local viral–viral or virus–host interactions. Epithelial cells within the oral cavity have been identified as a major site of EBV amplification, producing high viral loads in healthy subjects that cannot be explained solely by lytic replication within B cells (2). SARS-CoV-2 infection may lead to local inflammation and immune activation at mucosal surfaces that could disrupt mechanisms that normally constrain EBV lytic replication. Conversely, lytic EBV replication has been shown to enhance expression of the SARS-CoV-2 receptor ACE2 on epithelial cells and enhance SARS-CoV-2 entry(16). Together, these findings are consistent with a model in which SARS-CoV-2–driven mucosal immune perturbations and potentially co-infected cells, facilitate EBV reactivation at epithelial sites which in turn facilites SARS-CoV-2 dissemination, resulting in coordinated increases in both viruses within saliva. How this relationship may be altered by antiviral treatment, which in the case of Paxlovid can lead to SARS-CoV-2 rebound in up to 25% of individuals (13, 17), warrants further enquiry.

Notably, we did not observe an association between EBV salivary shedding, and the induction of a systemic EBV antibody responses measured on day 28 following SARS-CoV-2 infection. Rather, we found that individuals with no detectable EBV reactivation in saliva had increased EA-D IgG titers in blood compared to those with EBV reactivation, suggestive of a possible protective immune response. This dissociation highlights an important distinction between localized mucosal EBV activity and systemic EBV reactivation and suggests that salivary EBV measurements alone may be insufficient to detect reactivation dynamics at the systemic level. Accordingly, no association between EBV shedding in saliva and the frequency of latently infected peripheral memory B cell was observed in healthy adults(2). Given that peripheral memory B cells are a key reservoir of latent EBV and that the frequency of latently infected cells has been found to be stable over several months in the absence of pathology(2), it will be important to establish how this population may contribute to EBV DNAemia in blood and how this is modulated by COVID-19. This is particularly relevant in cases of Long COVID, which has been linked to systemic EBV reactivation(8, 18). Ultimately, our data provides a rationale for trials seeking to more aggressively control SARS-CoV-2 replication during the acute phase and for assessment of the effect of such strategies on mitigating EBV reactivation.

Our findings also have potential implications for oral and periodontal health. EBV has been implicated in the pathogenesis of periodontal disease, particularly through interactions with bacterial pathogens that can promote viral reactivation (19). Prolonged or repeated EBV reactivation in saliva during SARS-CoV-2 infection may therefore contribute to local inflammatory processes within the oral environment (20). While this study was not designed to evaluate clinical oral outcomes, our results provide a rationale for future investigations examining the impact of SARS-CoV-2–associated EBV reactivation on oral health.

This study has several limitations. The sample size was modest, which may have limited statistical power to detect smaller effect sizes and ability to adjust for potential confounders. Although the longitudinal design allowed for detailed characterization of within-host viral shedding dynamics, the observational nature of the study precludes causal inference regarding the relationship between SARS-CoV-2 infection and EBV reactivation. Additionally, systemic EBV antibody responses were assessed at a single convalescent time point, which may not capture dynamic changes occurring earlier or later in infection.

In conclusion, this study provides evidence that SARS-CoV-2 infection is associated with sustained salivary EBV shedding, that concurrent high levels of EBV and SARS-CoV-2 in saliva are correlated, but that salivary EBV shedding dynamics do not drive systemic EBV antibody responses. The findings correlating the EBV and SARS-CoV-2 levels extend prior inpatient observations to an outpatient environment but highlight an important discrepancy between EBV DNA shedding in the oral cavity and systemic EBV antibody responses. Further research with larger cohorts and mechanistic assays would be helpful to elucidate the biological pathway triggering systemic EBV antibody responses.

## Data Availability

All data produced in the present study are available upon reasonable request to the authors

## Author approval

All authors have seen and approved the manuscript.

